# AVAILABILITY, ACCESSIBILITY AND AFFORDABILITY OF COMPREHENSIVE CARE AMONG CHILDREN WITH CEREBRAL PALSY IN DAR ES SALAAM, TANZANIA: A CROSS SECTIONAL, ANALYTICAL HOSPITAL-BASED STUDY PROTOCOL (CAREGIVERS PERSPECTIVE)

**DOI:** 10.1101/2025.11.26.25341070

**Authors:** Ansaar Ismail Sangey, Florence Salvatory Kalabamu, Zameer Fakih, Francis Mchomvu, Zahra Morawej, Nasreen Ayoub Daud, Rhobi Gabriel Sisa

**Affiliations:** Resident Doctor, Dept of Paediatrics & Child Health – Kairuki University, Dar es Salaam Tanzania; Professor, Dept of Paediatrics & Child health – Kairuki University, Dar es Salaam Tanzania; Paediatrician, Paediatric Neurology clinic – Muhimbili National Hospital, Dar es Salaam Tanzania; Paediatrician, CCBRT Hospital – Dar es Salaam Tanzania; Psychiatrist, Kairuki University, Dar es Salaam Tanzania

**Keywords:** Cerebral Palsy, Comprehensive care, Availability, Affordability, Accessibility

## Abstract

**EXECUTIVE SUMMARY:** 

**Background:** Cerebral palsy is among the major causes of permanent disability in children worldwide. Whereas the overall global incidence rate of CP is 2.11 per 1000 live-births, in Africa incidence rate has been estimated at about 3.34 per 1000 live-births. Many caregivers and families of children with CP in African countries face multiple challenges that limit availability, accessibility and affordability of comprehensive care for children with Cerebral Palsy.

**Objectives:** To assess the availability, accessibility and affordability of comprehensive care among children with cerebral palsy attending specialized pediatric neurology clinics in Dar es Salaam, Tanzania.

**Design & Methods:** A cross-sectional, analytical hospital-based study will be conducted at two multi-specialty hospitals (Muhimbili National Hospital & CCBRT hospital) in Dar es Salaam, Tanzania from November 2025 – January 2026. Caregiver-child with CP pair will be the target population. Pre-tested questionnaire will be the main tool for data collection. A minimum of 229 children-caregiver pairs will be needed in order to attain a minimum study power of 80% given an α-level of 5%. Descriptive analysis will be conducted to assess the magnitude of availability, accessibility and affordability of care. A binary multivariable logistic regression model will be used to assess factors associated with availability, accessibility and affordability of care. Written informed consent will be obtained from caretakers of all study participants prior to recruitment into the study.

**Main outcomes measures:** Proportion of children-caregiver pairs with attained ‘comprehensive care’ for CP.

**STUDY’S STRENGHTS & LIMITATIONS:** *Strengths:*

- Study settings included multi-disciplinary health facilities for pediatric neurology care
- Inclusivity (i.e. caregiver-child pairs)
- Findings will be generalizable to the entire country since the 2 study settings receive patients from throughout the nation.

*Limitations:*

- Since the study is cross-sectional by design, temporal association (i.e. between exposure and outcomes) cannot be proven.
- Possibilities of recall biases from caregivers’ responses cannot be ruled-out.

## INTRODUCTION

Cerebral Palsy that refers to “a group of permanent disorders of movement and posture that cause activity limitations and are attributed to non-progressive disturbances that occurred in the developing fetal or infant brain” (1). They are usually accompanied by impairment in sensation, perception, cognitive function, communication, behavior, and may sometimes be associated with epilepsy and secondary musculoskeletal problems (1). Many caregivers and families of children with CP in African countries face multiple challenges that limit availability, accessibility and affordability of comprehensive care for children with Cerebral Palsy (2, 3). These challenges precipitate a significant setback in the management, outcomes and overall quality of life for children with CP (2, 3). This proposed study sets to assess availability, accessibility and affordability of comprehensive care among children with cerebral palsy from parents’/care takers’ perspectives.

The overall global incidence rate of CP is 2.11 per 1000 live births (4). In Africa incidence rate has been estimated at about 3.34 per 1000 live births (5). The African continental figure is also variable among countries. For instance, a study done in rural Uganda showed incidence of about 1·8–2·3 cases per 1000 children (6). In Tanzania few studies are available, and a study done in Mbeya revealed incidence of CP at about 17.77 per 1000 live births (7).

Comprehensive care refers to “*the coordinated delivery of the total health care required or requested by a patient*” (8). Children with CP face a myriad of complications including musculoskeletal complications such as spasticity, contractures, joint deformities, hip dislocation, scoliosis, osteopenia as well as fractures. They also face neurological manifestations. Comprehensive care for children with CP involves multispecialty collaboration including *pediatric neurologists, physiotherapists, orthopedic surgeons, occupational and speech therapists, psychologists, ophthalmologists, speech, use of assisted devices* as well as *family centered approach* (9). It aims at maximizing the child’s functional independence and quality of life (9). In order to ensure comprehensiveness in assessment of care among children with cerebral palsy, other associated aspects need to be taken into account, namely *availability of care services, accessibility* as well as *affordability*. At present, there is scarcity of retrievable literatures that analyze aspects of comprehensive care among children living with cerebral palsy in Tanzania (2).

Availability in health refers to presence of service domains and sub-domains when accessing care (2, 10). For instance, in a study done in Ghana by Oguntade and his colleagues, they found the most salient barriers at the super-structural, structural and environmental levels (3). Moreover, accessibility to healthcare refers to the ability of individuals to obtain timely, affordable, and appropriate healthcare services when the need arises (11). It involves removing barriers that may hinder people from seeking and achieving necessary medical care (11). However, until now – there are virtually no quantitative based data to substantiate availability, and accessibility of comprehensive care among children with cerebral palsy in African set ups.

Apart from accessibility, caretakers and children with cerebral palsy are also speculated to have challenges in affording required and timely healthcare services (2,3). Health care affordability refers to measure(s) of individual ability to pay for healthcare costs (12). It can be described as the balance between the costs of care and the financial means of the individuals or families needing care (12). Previous research reports have gone far wider than healthcare costs in childhood which include quality of life and especially how it affects the construct of adjusted life years (13–15). At present, there is strong evidence to suggest that childhood illnesses to have deleterious consequences that last entire human lifespan, with most consequences later in old age (16–18). Several scholars have already provided evidence suggesting higher rates of ageing process in not only sub-Saharan Africa (19, 20), but also Tanzania (21, 22). They all have philosophical and contextual origin that dates back to *David Barker’s hypothesis* that went public back in England sometimes in 1986 (23). Thus, this proposed study will address the gap in quantitative estimates on availability, accessibility and affordability of comprehensive care among children with cerebral palsy attending specialized paediatric neurology healthcare facilities in Dar es Salaam, Tanzania.

## METHODS AND ANALYSIS

### Study design

The study will be a cross-sectional hospital-based analysis. The rationale behind the suggested study design is based on the fact that it will mainly be focused on screening assessment of availability, accessibility and affordability of comprehensive care among children with cerebral palsy. Thus, both exposure (children with CP on comprehensive care) and outcomes (availability, affordability and accessibility) will be assessed at the same time and analyses their relationship using inferential statistics.

### Study settings

This study will be conducted in two major multispecialty hospitals namely Muhimbili National Hospital (MNH) and CCBRT Hospital both in Dar es Salaam, Tanzania. These two (2) hospitals have been selected because of their multi-specialty handling of different neurologic conditions for both children and adults in Dar es Salaam city. Besides, Muhimbili National Hospital is the national public tertiary referral hospital in Tanzania, with good mix of multi-disciplinary specialists’ team (24). Children with cerebral palsy are being served and attended at specialized outpatient pediatric neurology clinic (24). An average of 30 patients with CP are seen at the clinic per month.

On the other hand, CCBRT hospital acts as a specialized private rehabilitation center in Dar es Salaam, Tanzania (25). It provides rehabilitation services to both adults and pediatric patients. About 20000 children attend its pediatric units on an annual basis. It is also a well-known center for its services on children with CP. On average, 120 CP patients are being attended per month at the facility. It also conducts numerous outreach programs for children with CP.

### Study population

Children with cerebral palsy attending specialized pediatric neurology clinics in Dar es Salaam, Tanzania.

### Target population

Caregiver-child pair with cerebral palsy who attained comprehensive care attending Muhimbili National Hospital and CCBRT hospital in Dar es Salaam, Tanzania.

### Sampling method

Proportionate sampling will be used across the two hospitals. Criterion based, serial recruitment whereby all children who meet specified inclusion criteria will be sequentially enrolled until the targeted sample size is reached.

### Sample size and sample size estimation

Sample size will be based on infinite population assumption (prevalence = 50%) since there has been no available/retrievable studies on the matter in similar areas.

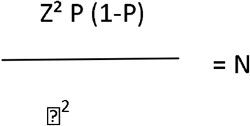

Whereby:

N = Minimum sample size needed

Z = Percentage point of normal distribution corresponding to the level of confidence. If the level of significance is 95% then Z is 1.96.

⍰ = margin of error i.e. 0.05.

P = Proportion of Children with CP (assumed as 50% since no available study is present)

Hence the minimum sample size will be

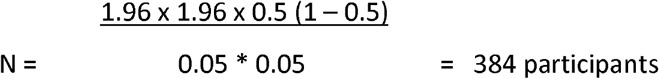

Adjusting for finite population, the minimum number of children (n’) needed to get the appropriate study power (at least 80%) will be given by the formula (26):

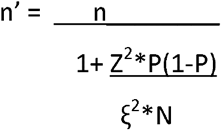

where by:

n’ = minimum sample size (adjusting for finite population)

n = minimum sample size from the Cochrane’s formula

Z = Standard Z-score

P = proportion of children with CP receiving comprehensive care (if unknown = 50%)

ξ = error rate (at alpha level of 5% = 0.05)

N = total population size = (30/month at MNH + 120/month at CCBRT) * duration of data collection (3 months)

Therefore,

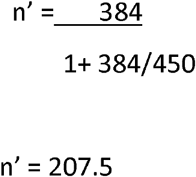

Therefore, the minimum number of caregivers-child pairs with CP needed to be studied in order for the study to get at least 80% power and considering an alpha level of 5% will be around 208.

Adding 10% non-response, the minimum sample size taken will be 229 Caregiver-child pair with Cerebral palsy attending MNH and CCBRT hospitals in Dar es Salaam. Specifically, about 46 caregiver-child pairs will be recruited from MNH while 183 caregivers-child pairs will be recruited from CCBRT hospitals.

### Eligibility criteria

All children with cerebral palsy from 0–18 years will be eligible to participate if caregivers- children pair with CP attending clinics at MNH and CCBRT hospitals, their diagnosis of CP made at least 6 months prior to recruitment date. However, Children with other neurodevelopmental disorders apart from CP, caregivers without ability to communicate in English and/or Kiswahili languages will be excluded from this proposed study.

### Study variables

#### Dependent (outcome) variable

Proportion of comprehensive care among children with CP, whereby comprehensive care will be a composite measure of the indices of availability, accessibility and affordability.

#### Independent (predictor) variables

**Socio-demographic factors** such as age (continuous) and sex (categorical) of the child, caregiver’s education level (categorical), caregivers’ relations to the child (categorical), caregivers’ highest level of formal schooling (categorical), total number of children in the family (discrete), type of family (categorical), child’s birth order (discrete).

**Healthcare-related factors** such as availability of comprehensive care CP, services and treatment available for comprehensive care, distance form household to hospital, time spent to reach hospital, time spent to access care in the hospital, clinic attendance pattern, factors affecting regular clinic attendance and health insurance coverage.

**Economic factors** such as affordability of comprehensive care, health insurance status (categorical), expenses spent in accessing the care, external financial support, economic and employment status of the care giver.

**Other factors** such as stigma caregivers face

### Data collection and tool

Data will be collected using a structured questionnaire via interview. It will comprise of demographic information of the child and the caregiver, type of CP the child is having, any comorbid conditions the child is suffering, the drugs, services and care the child attains in the hospital. Also factors which contribute and affect availability, accessibility and affordability of comprehensive care for children with CP will be collected. Data will be collected by the principal investigator and his trained assistants. Research assistants will be clinic nurses at Muhimbili National Hospital and CCBRT hospital in Dar es Salaam. Each interview will last about 20 minutes. Two versions of questionnaire will be prepared in both English and Kiswahili language for feasibility purpose of data collection. A one-day training on good clinical and research practice will be provided by the PI to all research team members, including research assistants.

### Tool pretesting

The study tool will also be pre-tested prior to commencement of the study. Specifically, about 10% of the study sample size will be recruited at a different centre (Amana hospital in Dar es Salaam) in order to assess the test-retest reliability as well as construct and face validity. The tool will be shared with 3 experts in research related to children disability at MNH and CCBRT to assess the contents and rate the validity, readability and the test understandability of variables in 10% of participants from other hospitals, then it will be revised accordingly before data collection.

### Data processing and analysis plan

All data that will be collected will be triple-entered in a pre-designed template using SPSS™ (SPSS-IBM, Chicago, USA) software version 23. Data cleaning (e.g., checking for missing value, errors in data entry) will be done at the end of each business day by the PI himself. Thereafter, the cleaned dataset will be stored until analysis time.

Data analysis will be done using Stata version 23.0. Numerical data will be summarized using median and inter-quartile range while categorical data will be summarized using frequency and proportion. Moreover, there will be exploratory data analysis, that will include data reduction methods (e.g. principal component analysis), specifically tailored for those open-ended questions. Availability, accessibility and affordability of comprehensive care among children with CP will be in frequency and respective percentages in response category in Likert’s scale responses. Specifically, 5-item Likert responses will be used. They will be categorized into the formats *always, sometimes, neutral, rarely and never*. Factors associated with availability, accessibility, and affordability of CP care will be assessed using binary logistic regression analysis. Crude Odd Ration and respective 95% confidence intervals for respective independent variables with be computed in univariate model. Factors with significance at p value of less or equal to 0.2 will be included in the multivariate model. Adjusted Odd Rations and respective 95% confidence intervals will be computed. Factors with p value of less or equal to 0.05 in a multivariate model will be considered to be independently associated with dependent variables (availability, accessibility, and affordability of CP care).

### Ethics and dissemination

Ethical clearance has been obtained from the Institutional Research Ethics Committee (IREC) of Kairuki University (KU). Also, permission will be obtained from hospital hierarchy of MNH and CCBRT hospital to conduct the study at their respective hospitals. Finally, those who will join the study will be asked to go through the informed consent and sign for the permission. The final written informed consent will be provided by the parent-care taker.

Informed consent will include the purpose of the study. Caretakers will be explained and assured that participation will not interfere care given at the hospital. They will also be informed that participation is voluntary and free to participate/withdraw from any part or whole questionnaire at any time. Besides, interviews will be conducted at a separate room/compartment in order to ensure maximum confidentiality between study participants and their caregivers on one side and the interviewer on another side. The questionnaire will not include the name of the patient nor the care givers. Information gathered during the study will be kept confidential and will not be disclosed to anyone apart from department and research staff. This is to preserve patients’ privacy and confidentiality.

## Data Availability

No data is available, this is a protocol research

## Authors’ contributions

AIS – Conceptualization, funding, data curation, data collection, drafting and reviewing and approving final draft

FSK – Conceptualization, data curation, supervising, drafting and reviewing and approving final draft

ZF Conceptualization, supervising and reviewing and approving final draft

FM – Conceptualization, supervising and reviewing and approving final draft

ZM Conceptualization, data curation, supervising, drafting and reviewing and approving final draft

NAD – Conceptualization, data curation, drafting and reviewing and approving final draft

RGS Conceptualization, data collection, drafting and reviewing and approving final draft

## Funding statement

No funds has been granted for this specific research study. Data collection will be a self - initiatives of the authors.

## Competing interests

All authors declare no competing interest exists in conceptualization of this proposed study nor will they declare any interest in data/findings thereof

## Notes

### Competing Interest Statement

The authors have declared no competing interest.

### Author Declarations

Institutional Research Ethics Committee of Kairuki University

